# Parental intention, attitudes, beliefs, trust and deliberation towards childhood vaccination in the Netherlands in 2022: Indications of change compared to 2013

**DOI:** 10.1101/2023.07.24.23291934

**Authors:** Marthe Knijff, Alies van Lier, Maartje Boer, Marion de Vries, Jeanne-Marie Hament, Hester E. de Melker

## Abstract

**Background and aim:** Vaccine uptake within the Dutch National Immunisation Programme (NIP) has slightly declined since the COVID-19 pandemic. We studied psychosocial factors of vaccine uptake, namely parental intention, attitudes, beliefs, trust and deliberation (i.e., self-evidence), before (2013) and two years into the pandemic (2022).

**Methods:** In 2022 and 2013, parents with a young child (aged <3.5 years) participated in online surveys on vaccination (n=1,000 and 800, (estimated) response=12.2% and 37.2%, respectively). Psychosocial factors were measured on 7-point Likert scales. Multivariate logistic regression analysis was used to study differences between parents in 2022 and 2013 in ‘negative’ scores (≤2) of psychosocial factors.

**Results:** In both 2022 and 2013, most parents with a young child expressed positive intention (2022=83.1%, 2013=87.0%), attitudes (3 items: 2022=66.7%-70.9%, 2013=62.1%-69.8%) and trust (2022=51.8%, 2013=52.0%) towards the NIP and felt that vaccinating their child was self-evident (2022=57.2%, 2013=67.3%). Compared to parents with a young child in 2013, parents with a young child in 2022 had significantly higher odds of reporting negative attitudes towards vaccination (3 items combined: OR=2.84), believing that vaccinations offer insufficient protection (OR=4.89), that the NIP is not beneficial for the protection of their child’s health (OR=2.23), that vaccinating their child does not necessarily protect the health of other children (OR=2.24) or adults (OR=2.22) and that vaccinations could cause severe side effects (OR=2.20), preferring natural infection over vaccination (OR=3.18) and reporting low trust towards the NIP (OR=1.73).

**Conclusions:** Although most parents had positive intention, attitudes and trust towards vaccination and perceived vaccinating their child as self-evident, proportions of parents with negative scores were slightly larger in 2022 compared to 2013. Monitoring these determinants of vaccine uptake and developing appropriate interventions could contribute to sustaining high vaccine uptake.

## 1 Background

Vaccination is considered one of the safest and most effective primary health care measures to prevent mortality and morbidity due to infectious diseases [1]. A high and homogeneous vaccine uptake is a key factor for success of vaccination programmes. Besides offering direct protection to vaccinated individuals, high vaccination coverage rates offer indirect protection to the overall community, or herd protection, by reducing transmission of infectious diseases [2, 3]. Participation in routine immunisation programmes, however, is not self-evident. During the COVID-19 pandemic several countries have reported perturbed continuation of routine infant vaccinations [4–6]. Vaccine uptake may be affected by health system barriers (e.g., health and language literacy barriers, accessibility barriers such as limited service hours and the geographical distance to the vaccination location and, in the case of the COVID-19 pandemic, barriers related to COVID-19 response measures). Next to such health system barriers, vaccine hesitancy among the public can form a major barrier to a high and homogenous vaccination uptake [7]. Vaccine hesitancy includes a wide range of attitudes and behaviours related to the reluctance or refusal to vaccinate despite the availability of vaccines [8].

In the Netherlands, children are offered vaccinations against 12 infectious diseases (diphtheria, hepatitis B, *Haemophilus influenzae* serotype b (Hib) disease, human papilloma virus (HPV) infection, measles, meningococcal ACWY disease, mumps, pneumococcal disease, polio, rubella, tetanus and pertussis) by the National Immunisation Programme (NIP). Vaccinations included in the NIP are administered on a voluntary basis and are free of charge [9]. Young children (i.e., aged ≤4 years) receive their vaccinations at child health clinics and older children (i.e., aged 9-14 years) are mainly vaccinated during group vaccinations. Vaccine uptake rates within the NIP have traditionally been high: most childhood vaccines have a coverage of around 90-95% [6]. The HPV vaccine for adolescent girls is an exception. From its introduction up till now, uptake of the HPV vaccine has steadily increased from 56% to nearly 70% [6]. Between reporting years 2015-2018, a small decrease in vaccination coverage was observed for infants born in 2012-2015 [6]. In reporting years 2020 and 2021, which predominantly covered vaccinations scheduled before the COVID-19 pandemic, uptake rates increased again [6]. Recently, however, it was shown that vaccine uptake of the first measles, mumps and rubella (MMR) vaccination, routinely scheduled at 14 months of age, declined during the first months of the COVID-19 pandemic. Initially, uptake dropped by 6-14% among infants born in 2019 who were scheduled for vaccination in March-September 2020 compared with the previous year. After catch-up vaccination, uptake still remained 1-2% lower [6, 10]. Figures on participation in the NIP for reporting year 2023 have shown that vaccine uptake of the first MMR vaccine has once more declined for infants born in 2020 [6]. In addition, there have been indications that vaccine uptake of the group vaccinations scheduled during the spring of 2022 in particular has been falling behind. Similar trends have been observed for other vaccinations scheduled during the COVID-19 pandemic [6].

Vaccine hesitancy towards COVID-19 vaccines [11, 12] might have influenced parental attitudes and perceptions towards routine childhood vaccination. Available European studies have largely focussed on the general public and vaccination in general rather than parents of children eligible for routine vaccination programmes [13, 14]. A report published by the European Commission revealed that across the European Union, the public’s overall vaccine confidence (i.e., perceptions towards the importance, safety, and effectiveness of vaccination) has significantly declined since the beginning of 2020. For the Netherlands, they reported year-on-year declines in confidence towards general vaccination between 2018, 2020 and 2022 [13]. Similarly, a study conducted in the United Kingdom showed that attitudes towards the safety, effectiveness, necessity of vaccination have significantly decreased among the general public since the onset of the pandemic [14]. Similar observations were made in a report recently published by the United Nations Children’s Fund (UNICEF). In 52 out of 55 countries studied worldwide, the public’s perception of the importance of vaccines for children declined during the COVID-19 pandemic. Perceptions of vaccine safety and effectiveness also showed declines, but not as pronounced [15]. Several studies from the United States of America (USA) have suggested that parental hesitancy and concerns about childhood vaccination increased somewhat during the COVID-19 pandemic [16–18]. Dutch child health clinic professionals have indicated that parents exhibited signs of decreased trust towards the NIP and “vaccine fatigue” [19] during the COVID-19 pandemic.

According to the Theory of Planned Behaviour [20], behaviour (i.e., vaccine uptake in the case of vaccination behaviour) is most strongly predicted by intention (i.e., the willingness to vaccinate). In a study conducted by Harmsen [21] in 2013, they developed a theory- and data-based model of psychosocial determinants of parents’ intention to vaccinate their child. Their model suggested that parental intentions to vaccinate their child were best predicted by a positive attitude, having trust in the NIP, high anticipated regret of not vaccinating, low perceived barriers of getting their child vaccinated, not making a deliberate decision (i.e., low deliberation) and having positive beliefs about vaccines. Positive beliefs about vaccines, high moral norms about vaccination and high trust in the NIP had the strongest influence on parents’ attitudes towards childhood vaccination. They largely based their model on a study by Paulussen et al. [22], who examined determinants of Dutch parents’ vaccination decisions and based their theoretical framework on the Theory of Planned Behaviour [20] and the Social Cognitive Theory [23].

In this study, we aimed to build upon previous work from Harmsen [21]. To ensure feasibility and conciseness of the study, we made an expert-based selection of a subset of the aforementioned psychosocial factors of parents’ vaccine uptake believed to be most relevant. Our main objective was to gain more insight into parental intention, attitudes, beliefs, trust and deliberation towards the current NIP and assess how these factors may have changed in 2022 compared to 2013. In addition, we assessed parents’ willingness towards adopting potential future expansions of the NIP (i.e., additional vaccines) and whether this willingness differed between 2022 and 2013. Finally, we investigated whether there were differences in the studied psychosocial factors of vaccine uptake between parents with a young child (aged <3.5 years) and parents with an older child (aged 9-14 years) in 2022, as there were indications that vaccine uptake of the group vaccinations (which are given to older children, i.e., aged 9-14 years) in particular had decreased in 2022.

## 2 Methods

### 2.1 Study population and design

Two cross-sectional surveys with similar design were carried out in 2022 and 2013 [21]. Participants were selected and invited in collaboration with [Flycatcher Internet Research](https://www.flycatcher.eu/en), a Dutch ISO-certified research agency. Flycatcher has their own online panel, but also works together with several partner agencies. The Flycatcher panel consists of more than 10,000 Dutch participants aged ≥18 years. Panel participants have voluntarily and actively registered their consent to participate in surveys via double opt-in.

According to Dutch law (i.e., the Medical Research Involving Human Subjects Act (WMO)), the nature of these general internet-based surveys among healthy volunteers does not require formal medical ethical approval (www.ccmo.nl).

#### 2022 survey

In 2022, participants were selected from panels of Flycatcher’s partner agencies and predominantly invited to participate via email^1^ with a link to an online survey on their opinion about vaccination. Two groups of panel participants were invited on July 1^st^, 2022: (1) parents with at least one child aged <3.5 years (young child) and (2) parents with at least one child aged 9-14 years (older child). Parents had access to the questionnaires until at least a 1,000 parents per group had completed the questionnaire.

Parents with a young child had access to the questionnaire until July 5^th^, 2022. The total number of approached parents with a young child was estimated^1^ to be approximately 8,200 (estimated response: 12.2%). Parents with an older child had access to the questionnaire until July 8^th^, 2022. In total, 4,779 parents with an older child were approached^2^ and a reminder was sent to 806 low-educated parents who had not (fully) completed the questionnaire to improve representativeness of the sample (response: 20.9%).

Before they were sent out, Flycatcher pre-tested the questionnaires to detect potential technical errors and to check whether questions were understood. In order to improve representativeness of the samples, Flycatcher stratified samples by gender and educational level and weighted data for gender and educational level based on population statistics obtained from Statistics Netherlands.

#### 2013 survey

For the survey conducted in 2013, Flycatcher approached a total of 2,150 parents with a young child from their own panel and a panel from another agency in order to achieve a sufficient percentage of immigrant parents. In 2013, no parents of older children were included. The 2013 survey was completed by 800 parents with a young child (response: 37.2%). Similar procedures were performed in terms of stratification and weighting as described for the 2022 survey [21].

### 2.2 Questionnaire

Supplemental Table 1 provides an overview of the questions that were part of the 2022 survey and the corresponding questions from the 2013 survey. Surveys were designed by researchers from the National Institute for Public Health and the Environment in consultation with Flycatcher. Questions covered various topics, such as demographic information of the participants, self-reported vaccine uptake, psychosocial factors of vaccine uptake and participants’ opinions on the NIP. Questions were mostly similar across the years; some small alterations were made to a selection of the questions from the 2013 survey for the 2022 survey (Supplemental Table 1).

### 2.3 Outcomes

Self-reported *vaccine uptake* was measured by asking parents whether their child participated in the NIP (answer categories: ‘yes’, ‘partially’ and ‘no’).

The selected psychosocial factors of vaccine uptake were based on previous work by Harmsen [21], in which they developed a model of determinants of parents’ vaccination intention. All items from these factors were measured on 7-point Likert scales. Parents’ *intention* to vaccinate their child according to the NIP was measured using a single item. Parental *attitudes* towards the NIP were measured using three items and concerned their perception of the value (good vs. bad), importance and necessity of vaccination. For attitude, a composite measure was constructed by averaging the three single-item scores, which was used as an outcome in multivariate analysis (see section 3.4). The Cronbach’s alpha was 0.94, indicating good internal reliability of the attitude scale [24]. Seven items on *beliefs about vaccination* were used, which measured parents’ perceptions about the effectiveness, safety and side effects of vaccination. Parents’ *beliefs about infectious diseases* (i.e., preference of natural infection over vaccination), *deliberation* about their decision to vaccinate (i.e., whether they considered vaccinating their child to be self-evident) and *trust* towards the NIP were all measured with single items. Details regarding these items and factors can be found in Supplemental Table 1.

We also assessed parents’ *intention towards potential future expansions of the NIP* (Supplemental Table 1). For this factor 5 items were used, which were measured on 7-point Likert scales as well. Parents were asked whether they would be willing to vaccinate their child against rotavirus infection, chickenpox, respiratory syncytial virus (RSV) infection, meningococcal B disease and influenza if vaccinations against these infectious diseases were to be incorporated into the NIP. In 2022, these questions were only presented to parents with a young child, as they were less relevant for parents with an older child (i.e., these vaccinations are offered to young children).

For all individual psychosocial items, parents were grouped into ‘negative’ (score 1-2), ‘less pronounced’ (score 3-5) and ‘positive’ (score 6-7). Items were also analysed in the continuous scale (range 1-7) and reverse-coded when applicable.

### 2.4 Statistical analysis

Descriptive analysis (frequencies and measures of central tendency, i.e., mean and median, and variability, i.e., standard deviation and interquartile range) was used to describe demographic characteristics of the unweighted and weighted samples and psychosocial factors of the weighted samples only. Demographic differences between parents with a young child in 2013 and 2022 were analysed with two-sample t-tests for age and univariate chi-square tests for all other demographic characteristics. Mann-Whitney U tests were used to compare psychosocial factors in the continuous scale (range 1-7) between parents with a young child in 2022 and 2013. To assess whether parents with a young child had become more negative and/or positive towards vaccination in 2022 compared to 2013, scores of psychosocial factors were dichotomised into ‘negative’ (score ≤2; range 1-7) vs. ‘other’ (score >2) and ‘positive’ (score ≥6) vs. ‘other’ (score <6) and analysed with univariate chi-square tests. We used multivariate logistic regression analysis to control for demographic characteristics of parents with a young child in 2022 and 2013. Hereby, we restricted the analysis to the negative variants of the dichotomised psychosocial factors – ‘negative’ (score ≤2) vs. ‘other’ (score >2). Models were adjusted for parents’ age, income level and country of birth. The same approach was taken for analysis of differences between parents with a young child and parents with an older child in 2022.

Except for descriptive analysis of demographic characteristics, analyses were conducted using weighted data only and were considered statistically significant if *p*-values were below 0.05. Analyses were conducted in R 4.2.0 [25] and RStudio 2022.2.2.485 [26] using the survey package [27].

### 2.5 Sensitivity analysis

A sensitivity analysis was performed to see whether results were robust to using different cut-offs for the psychosocial factors of vaccine uptake. In the univariate analysis, psychosocial factors were dichotomised into ‘negative’ (score ≤3; range 1-7) vs. ‘other’ (score >3) and ‘positive’ (score ≥5) vs. ‘other’ (score <5). Here too, the multivariate analysis was restricted to the negative variants of the dichotomised psychosocial factors – in this case ‘negative’ (score ≤3) vs. ‘other’ (score >3).

## 3 Results

### 3.1 Demographic characteristics

#### Parents with a young child

Demographic characteristics of both the unweighted and weighted samples are presented in Table 1. After weighting for sex and educational level, parents with a young child from 2022 were shown to be slightly but significantly older than parents from 2013. In addition, in 2022 significantly more parents had a household income below average and significantly less parents did not know their household income level or preferred to withhold from answering. Furthermore, in the 2022 sample, significantly more parents were born in the Netherlands. No significant differences were observed in terms of sex, educational level and region of residence.

**Table 1.** Demographic characteristics of the unweighted and weighted samples of parents in 2013 and 2022. Weighting was performed for sex and educational level.

|  | 2013 young child |  | 2022 young child |  | 2022 older child |  | *p*-values<sup>1</sup> |  |
| --- | --- | --- | --- | --- | --- | --- | --- | --- |
|  | Unweighted,<br>*n* = 800 | Weighted,<br>*n* = 797 | Unweighted,<br>*n* = 1000 | Weighted,<br>*n* = 998 | Unweighted,<br>*n* = 1000 | Weighted,<br>*n* = 1002 | 2013 vs.<br>2022 young<br>child | 2022 young<br>vs. older<br>child |
| Age<sup>2</sup>, mean (SD) | 33.0 (3.8) | 32.9 (3.9) | 35.1 (5.5) | 35.4 (5.6) | 45.1 (6.6) | 45.2 (6.6) | **<0.001** | **<0.001** |
| Sex |  |  |  |  |  |  | 0.6 | >0.9 |
| Female | 70.4% | 51.8% | 59.6% | 49.9% | 54.2% | 49.8% |  |  |
| Male | 29.6% | 48.2% | 40.1% | 49.8% | 45.7% | 50.1% |  |  |
| Other<sup>3</sup> | 0.0% | 0.0% | 0.3% | 0.4% | 0.1% | 0.1% |  |  |
| Educational level<sup>4</sup> |  |  |  |  |  |  | 0.8 | 0.9 |
| Low | 6.6% | 20.9% | 9.8% | 19.1% | 15.6% | 18.5% |  |  |
| Medium | 31.8% | 44.2% | 47.8% | 44.5% | 51.4% | 45.6% |  |  |
| High | 61.6% | 34.9% | 42.4% | 36.4% | 33.0% | 36.0% |  |  |
| Household income level<sup>5</sup> |  |  |  |  |  |  | **0.001** | **<0.001** |
| Below average<br>(<€36.500) | 6.8% | 10.5% | 15.8% | 17.8% | 14.5% | 14.1% |  |  |
| Average (€36.500-<br>€43.500) | 19.5% | 21.3% | 20.1% | 20.1% | 14.7% | 14.4% |  |  |
| Above average<br>(≥€43.500) | 54.6% | 49.2% | 50.7% | 49.1% | 45.4% | 47.1% |  |  |
| Unknown/prefer not to<br>say | 19.1% | 19.0% | 13.4% | 13.1% | 25.4% | 24.3% |  |  |
| Country of birth |  |  |  |  |  |  | **<0.001** | 0.2 |
| The Netherlands | 87.5% | 81.4% | 96.0% | 95.7% | 96.8% | 96.8% |  |  |
| Other | 12.5% | 18.6% | 4.0% | 4.3% | 3.2% | 3.2% |  |  |
| Other parent's/guardian's<br>country of birth |  |  |  |  |  |  | **<0.001** | 0.3 |
| The Netherlands | 85.9% | 83.8% | 93.7% | 93.6% | 94.8% | 94.8% |  |  |
| Other | 13.9% | 15.8% | 5.8% | 6.0% | 4.5% | 4.6% |  |  |

|  | 2013 young child |  | 2022 young child |  | 2022 older child |  | *p*-values<sup>1</sup> |  |
| --- | --- | --- | --- | --- | --- | --- | --- | --- |
|  | Unweighted,<br>*n* = 800 | Weighted,<br>*n* = 797 | Unweighted,<br>*n* = 1000 | Weighted,<br>*n* = 998 | Unweighted,<br>*n* = 1000 | Weighted,<br>*n* = 1002 | 2013 vs.<br>2022 young<br>child | 2022 young<br>vs. older<br>child |
| Unknown/not applicable | 0.2% | 0.4% | 0.5% | 0.5% | 0.7% | 0.6% |  |  |
| Region of residence<sup>6</sup> |  |  |  |  |  |  | 0.051 | 0.8 |
| North-East | 28.3% | 28.7% | 31.2% | 31.3% | 33.1% | 32.6% |  |  |
| West | 39.9% | 40.9% | 44.2% | 44.5% | 42.4% | 43.1% |  |  |
| South | 31.8% | 30.4% | 24.6% | 24.1% | 24.5% | 24.3% |  |  |

Abbreviation: SD, standard deviation.
<sup>1</sup>A two-sample t-test for age and chi-square tests for the other demographic characteristics were performed on data weighted for sex and educational level. *P*-values below <0.05 were considered significant (bold and underlined in table).
<sup>2</sup>Parents' age ranged from 25-40 years in 2013 and 20-60 years in 2022.
<sup>3</sup>In the 2022 survey, there was an additional answer option ('other') for the question concerning sex. In order to ensure comparability of the surveys, these data were not included in the corresponding chi-square analyses.
<sup>4</sup>Educational level was grouped as: low (none, primary, lower vocational or lower secondary education), medium (secondary vocational or higher secondary education), high (higher professional or university education)
<sup>5</sup>In 2013, different income cut-offs were used to indicate the different categories: below average (<€23.000), average (€23.000-€34.000), above average (≥€34.000), don't know/prefer not to say.
<sup>6</sup>Parents were grouped into regions according to the Department for Vaccine Supply and Prevention Programmes (DVP) regions: North-East (Groningen, Friesland, Drenthe, Overijssel, Flevoland and Gelderland), West (Utrecht, Noord-Holland and Zuid-Holland), South (Noord-Brabant, Limburg and Zeeland). Among parents with a young child in 2013, there was one parent for whom information about their province of residence was not available. These data are not included in the table and corresponding chi-square analyses.

#### Parents with an older child

Parents with an older child only differed significantly from parents with a young child in 2022 with regard to age and household income level (Table 1). Parents with an older child were significantly older than parents with a young child in 2022. Additionally, significantly less parents with an older child had an income below average or an average income and significantly more of them did not know their income or preferred to withhold from answering.

### 3.2 Vaccine uptake, intention, attitudes, beliefs, trust and deliberation towards the current NIP

#### Parents with a young child

In general, the majority of parents with a young child indicated that their child participated in the NIP, expressed positive intention, attitudes, and trust towards the NIP and felt that vaccinating their child was self-evident in both 2022 and 2013 (Table 2).

**Table 2.** Overview and univariate comparison of vaccine uptake, intention, attitudes, beliefs, trust and deliberation towards the current NIP and intention towards potential expansions of the NIP among parents in 2013 and 2022. Data were weighted for sex and educational level.

|  | 2013 | 2022 |  | *p*-values<sup>1</sup> |  |
| --- | --- | --- | --- | --- | --- |
|  | Young child,<br>*n* = 797 | Young child,<br>*n* = 998 | Older child,<br>*n* = 1002 | 2013<br>vs.<br>2022<br>young<br>child | 2022<br>young<br>child<br>vs.<br>older<br>child |
| *Vaccine uptake*<sup>2</sup> |  |  |  |  |  |
| Participation of (youngest) child within the NIP. |  |  |  |  |  |
| No | 2.6% | 5.4% | 4.2% | **0.011** | 0.3 |
| Partial | 6.9% | 6.6% | 9.3% |  |  |
| Yes | 90.5% | 88.0% | 86.5% | 0.2 | 0.3 |
| *Intention*<sup>2</sup> |  |  |  |  |  |
| Intention to vaccinate (youngest) child according to the NIP in the future. |  |  |  |  |  |
| Negative (score 1-2) | 4.4% | 6.1% | 6.8% | 0.2 | 0.5 |
| Less pronounced (score 3-5) | 8.6% | 10.8% | 12.4% |  |  |
| Positive (score 6-7) | 87.0% | 83.1% | 80.8% | 0.080 | 0.2 |
| Median [IQR] | 7.0 [6.0, 7.0] | 7.0 [6.0, 7.0] | 7.0 [6.0, 7.0] | 0.6 | **0.047** |
| Mean (SD) | 6.3 (1.4) | 6.2 (1.5) | 6.1 (1.6) |  |  |
| *Attitude* |  |  |  |  |  |

|  | 2013 | 2022 |  | *p*-values<sup>1</sup> |  |
| --- | --- | --- | --- | --- | --- |
|  | Young child,<br>*n* = 797 | Young child,<br>*n* = 998 | Older child,<br>*n* = 1002 | 2013<br>vs.<br>2022<br>young<br>child | 2022<br>young<br>child<br>vs.<br>older<br>child |
| I think vaccinating my child(ren)<br>according to the NIP is: very bad<br>– very good |  |  |  |  |  |
| Negative (score 1-2) | 1.4% | 4.5% | 3.8% | **<u>0.002</u>** | 0.5 |
| Less pronounced (score 3-5) | 28.8% | 25.1% | 22.2% |  |  |
| Positive (score 6-7) | 69.8% | 70.5% | 74.0% | 0.8 | 0.093 |
| Median [IQR] | 6.0 [5.0, 7.0] | 6.0 [5.0, 7.0] | 6.0 [5.0, 7.0] | 0.3 | 0.2 |
| Mean (SD) | 5.9 (1.2) | 5.8 (1.4) | 5.9 (1.4) |  |  |
| I think vaccinating my child(ren)<br>according to the NIP is: very<br>unimportant – very important |  |  |  |  |  |
| Negative (score 1-2) | 2.2% | 4.9% | 4.5% | **<u>0.046</u>** | 0.7 |
| Less pronounced (score 3-5) | 28.4% | 24.2% | 21.0% |  |  |
| Positive (score 6-7) | 69.5% | 70.9% | 74.5% | 0.6 | 0.087 |
| Median [IQR] | 6.0 [5.0, 7.0] | 6.0 [5.0, 7.0] | 6.0 [5.0, 7.0] | 0.3 | 0.2 |
| Mean (SD) | 5.9 (1.2) | 5.9 (1.5) | 5.9 (1.4) |  |  |
| I think vaccinating my child(ren)<br>according to the NIP is: very<br>unnecessary – very necessary |  |  |  |  |  |
| Negative (score 1-2) | 3.3% | 5.9% | 6.5% | **<u>0.024</u>** | 0.6 |
| Less pronounced (score 3-5) | 34.6% | 27.4% | 25.2% |  |  |
| Positive (score 6-7) | 62.1% | 66.7% | 68.3% | 0.10 | 0.5 |
| Median [IQR] | 6.0 [5.0, 7.0] | 6.0 [5.0, 7.0] | 6.0 [5.0, 7.0] | **<u>0.048</u>** | 0.6 |
| Mean (SD) | 5.6 (1.3) | 5.7 (1.6) | 5.7 (1.6) |  |  |
| *Beliefs vaccination*<sup>3</sup> |  |  |  |  |  |
| Vaccinations offer insufficient<br>protection against the infectious<br>diseases they are targeting.* |  |  |  |  |  |
| Negative (score 1-2) | 4.0% | 16.5% | 17.7% | **<u><0.001</u>** | 0.5 |
| Less pronounced (score 3-5) | 52.7% | 43.4% | 35.0% |  |  |
| Positive (score 6-7) | 43.3% | 40.1% | 47.3% | 0.3 | **<u>0.002</u>** |
| Median [IQR] | 5.0 [4.0, 6.0] | 5.0 [3.0, 6.0] | 5.0 [3.0, 6.0] | **<u><0.001</u>** | **<u>0.005</u>** |
| Mean (SD) | 5.0 (1.3) | 4.5 (1.8) | 4.8 (1.9) |  |  |

|  | 2013 | 2022 |  | *p*-values<sup>1</sup> |  |
| --- | --- | --- | --- | --- | --- |
|  | Young child,<br>*n* = 797 | Young child,<br>*n* = 998 | Older child,<br>*n* = 1002 | 2013<br>vs.<br>2022<br>young<br>child | 2022<br>young<br>child<br>vs.<br>older<br>child |
| There are substances in vaccines that could be harmful to the health of my child(ren).* |  |  |  |  |  |
| Negative (score 1-2) | 10.5% | 12.6% | 13.0% | 0.2 | 0.8 |
| Less pronounced (score 3-5) | 76.1% | 55.0% | 49.7% |  |  |
| Positive (score 6-7) | 13.4% | 32.5% | 37.3% | **<0.001** | **0.028** |
| Median [IQR] | 4.0 [3.0, 4.0] | 4.0 [3.0, 6.0] | 5.0 [4.0, 6.0] | **<0.001** | **0.039** |
| Mean (SD) | 3.9 (1.3) | 4.5 (1.7) | 4.6 (1.7) |  |  |
| Vaccinations could lead to severe side effects later in life.* |  |  |  |  |  |
| Negative (score 1-2) | 5.4% | 11.1% | 11.3% | **<0.001** | 0.9 |
| Less pronounced (score 3-5) | 71.6% | 47.4% | 48.1% |  |  |
| Positive (score 6-7) | 23.0% | 41.5% | 40.5% | **<0.001** | 0.7 |
| Median [IQR] | 4.0 [4.0, 5.0] | 5.0 [4.0, 6.0] | 5.0 [4.0, 6.0] | **<0.001** | 0.6 |
| Mean (SD) | 4.4 (1.2) | 4.8 (1.7) | 4.8 (1.7) |  |  |
| The NIP is beneficial for the protection of the health of my child(ren). |  |  |  |  |  |
| Negative (score 1-2) | 2.1% | 5.0% | 4.2% | **0.002** | 0.4 |
| Less pronounced (score 3-5) | 39.5% | 40.0% | 37.2% |  |  |
| Positive (score 6-7) | 58.5% | 55.0% | 58.6% | 0.2 | 0.12 |
| Median [IQR] | 6.0 [5.0, 6.0] | 6.0 [5.0, 7.0] | 6.0 [5.0, 7.0] | 0.7 | 0.2 |
| Mean (SD) | 5.5 (1.1) | 5.4 (1.4) | 5.5 (1.4) |  |  |
| Vaccinating my child(ren) is a good way to protect the health of other children. |  |  |  |  |  |
| Negative (score 1-2) | 2.5% | 5.4% | 5.9% | **0.010** | 0.6 |
| Less pronounced (score 3-5) | 38.9% | 40.0% | 37.6% |  |  |
| Positive (score 6-7) | 58.6% | 54.6% | 56.5% | 0.2 | 0.4 |
| Median [IQR] | 6.0 [5.0, 6.0] | 6.0 [5.0, 7.0] | 6.0 [5.0, 6.0] | 0.2 | 0.5 |
| Mean (SD) | 5.4 (1.2) | 5.4 (1.5) | 5.4 (1.5) |  |  |
| Vaccinating my child(ren) is a good way to protect the health of adults. |  |  |  |  |  |
| Negative (score 1-2) | 4.3% | 8.8% | 7.7% | **0.001** | 0.4 |
| Less pronounced (score 3-5) | 52.3% | 44.1% | 43.7% |  |  |

|  | 2013 | 2022 |  | *p*-values<sup>1</sup> |  |
| --- | --- | --- | --- | --- | --- |
|  | Young child,<br>*n* = 797 | Young child,<br>*n* = 998 | Older child,<br>*n* = 1002 | 2013<br>vs.<br>2022<br>young<br>child | 2022<br>young<br>child<br>vs.<br>older<br>child |
| Positive (score 6-7) | 43.4% | 47.2% | 48.6% | 0.2 | 0.5 |
| Median [IQR] | 5.0 [4.0, 6.0] | 5.0 [4.0, 6.0] | 5.0 [4.0, 6.0] | 0.056 | 0.5 |
| Mean (SD) | 5.0 (1.3) | 5.1 (1.6) | 5.1 (1.6) |  |  |
| Vaccinating is a good way to protect against severe complications of infectious diseases. |  |  |  |  |  |
| Negative (score 1-2) | 3.6% | 4.3% | 3.8% | 0.5 | 0.6 |
| Less pronounced (score 3-5) | 46.2% | 37.7% | 33.8% |  |  |
| Positive (score 6-7) | 50.2% | 58.0% | 62.3% | **0.007** | 0.056 |
| Median [IQR] | 6.0 [4.0, 6.0] | 6.0 [5.0, 7.0] | 6.0 [5.0, 7.0] | **<0.001** | 0.3 |
| Mean (SD) | 5.2 (1.2) | 5.5 (1.4) | 5.6 (1.3) |  |  |
| *Beliefs disease* |  |  |  |  |  |
| Experiencing infectious diseases leads to a better and longer protection than a vaccination.* |  |  |  |  |  |
| Negative (score 1-2) | 8.3% | 23.8% | 19.7% | **<0.001** | **0.032** |
| Less pronounced (score 3-5) | 64.7% | 59.1% | 62.7% |  |  |
| Positive (score 6-7) | 27.0% | 17.1% | 17.6% | **<0.001** | 0.8 |
| Median [IQR] | 4.0 [3.0, 6.0] | 4.0 [3.0, 5.0] | 4.0 [3.0, 5.0] | **<0.001** | 0.2 |
| Mean (SD) | 4.4 (1.4) | 3.8 (1.7) | 3.9 (1.6) |  |  |
| *Trust* |  |  |  |  |  |
| When the government recommends vaccinations, I trust this is beneficial for my child(ren). |  |  |  |  |  |
| Negative (score 1-2) | 5.2% | 8.3% | 6.9% | **0.019** | 0.3 |
| Less pronounced (score 3-5) | 42.9% | 39.9% | 38.9% |  |  |
| Positive (score 6-7) | 52.0% | 51.8% | 54.2% | >0.9 | 0.3 |
| Median [IQR] | 6.0 [4.0, 6.0] | 6.0 [4.0, 7.0] | 6.0 [4.0, 6.0] | **0.022** | 0.6 |
| Mean (SD) | 5.2 (1.3) | 5.2 (1.7) | 5.3 (1.6) |  |  |
| *Deliberation* |  |  |  |  |  |
| Vaccinating my child(ren) is something I find self-evident. |  |  |  |  |  |
| Negative (score 1-2) | 6.0% | 7.7% | 7.1% | 0.2 | 0.6 |

|  | 2013 | 2022 |  | *p*-values<sup>1</sup> |  |
| --- | --- | --- | --- | --- | --- |
|  | Young child,<br>*n* = 797 | Young child,<br>*n* = 998 | Older child,<br>*n* = 1002 | 2013<br>vs.<br>2022<br>young<br>child | 2022<br>young<br>child<br>vs.<br>older<br>child |
| Less pronounced (score 3-5) | 26.7% | 35.1% | 36.7% |  |  |
| Positive (score 6-7) | 67.3% | 57.2% | 56.2% | **<0.001** | 0.6 |
| Median [IQR] | 6.0 [5.0, 7.0] | 6.0 [5.0, 7.0] | 6.0 [5.0, 7.0] | 0.061 | 0.4 |
| Mean (SD) | 5.6 (1.5) | 5.4 (1.6) | 5.4 (1.6) |  |  |
| *Intention potential NIP expansions* |  |  |  |  |  |
| Intention to vaccinate child against rotavirus infection. |  |  |  |  |  |
| Negative (score 1-2) | 17.5% | 10.9% | — | **<0.001** | — |
| Less pronounced (score 3-5) | 54.7% | 48.6% | — |  | — |
| Positive (score 6-7) | 27.8% | 40.5% | — | **<0.001** | — |
| Median [IQR] | 5.0 [3.0, 6.0] | 5.0 [4.0, 6.0] | — | **<0.001** | — |
| Mean (SD) | 4.4 (1.7) | 4.9 (1.7) | — |  | — |
| Intention to vaccinate child against chickenpox. |  |  |  |  |  |
| Negative (score 1-2) | 27.9% | 19.5% | — | **<0.001** | — |
| Less pronounced (score 3-5) | 38.6% | 40.8% | — |  | — |
| Positive (score 6-7) | 33.5% | 39.7% | — | **0.031** | — |
| Median [IQR] | 4.0 [2.0, 6.0] | 5.0 [3.0, 6.0] | — | **<0.001** | — |
| Mean (SD) | 4.2 (1.9) | 4.6 (2.0) | — |  | — |
| Intention to vaccinate child against RSV infection. |  |  |  |  |  |
| Negative (score 1-2) | 7.7% | 7.9% | — | 0.9 | — |
| Less pronounced (score 3-5) | 50.2% | 37.8% | — |  | — |
| Positive (score 6-7) | 42.1% | 54.3% | — | **<0.001** | — |
| Median [IQR] | 5.0 [4.0, 6.0] | 6.0 [4.0, 7.0] | — | **<0.001** | — |
| Mean (SD) | 5.0 (1.5) | 5.3 (1.6) | — |  | — |
| Intention to vaccinate child against meningococcal B disease. |  |  |  |  |  |
| Negative (score 1-2) | 4.6% | 4.6% | — | >0.9 | — |
| Less pronounced (score 3-5) | 48.3% | 31.3% | — |  | — |
| Positive (score 6-7) | 47.1% | 64.2% | — | **<0.001** | — |
| Median [IQR] | 5.0 [4.0, 6.0] | 6.0 [5.0, 7.0] | — | **<0.001** | — |
| Mean (SD) | 5.2 (1.4) | 5.7 (1.5) | — |  | — |

|  | 2013 | 2022 |  | *p*-values<sup>1</sup> |  |
| --- | --- | --- | --- | --- | --- |
|  | Young child,<br>*n* = 797 | Young child,<br>*n* = 998 | Older child,<br>*n* = 1002 | 2013<br>vs.<br>2022<br>young<br>child | 2022<br>young<br>child<br>vs.<br>older<br>child |
| Intention to vaccinate child against influenza. |  |  |  |  |  |
| Negative (score 1-2) | 44.7% | 36.9% | — | **<u>0.005</u>** | — |
| Less pronounced (score 3-5) | 43.5% | 44.9% | — |  | — |
| Positive (score 6-7) | 11.8% | 18.2% | — | **<u>0.002</u>** | — |
| Median [IQR] | 3.0 [2.0, 5.0] | 4.0 [2.0, 5.0] | — | **<u>0.034</u>** | — |
| Mean (SD) | 3.2 (1.8) | 3.5 (1.9) | — |  | — |
| Intention to vaccinate child against hepatitis A. |  |  |  |  |  |
| Negative (score 1-2) | — | 7.5% | — | — | — |
| Less pronounced (score 3-5) | — | 40.3% | — | — | — |
| Positive (score 6-7) | — | 52.2% | — | — | — |
| Median [IQR] | — | 6.0 [4.0, 7.0] | — | — | — |
| Mean (SD) | — | 5.3 (1.6) | — | — | — |

Abbreviations: IQR, interquartile range; SD, standard deviation; RSV, respiratory syncytial virus.
\*Items marked with an asterisk were reverse-coded. This means that parents grouped as ‘negative’ (score 1-2) agreed with the statement and parents grouped as ‘positive’ (score 6-7) disagreed with the statement.
<sup>1</sup>Mann-Whitney U tests were used to compare psychosocial factors in the continuous scale (range 1-7); chi-square tests were used to compare negative (vs. less pronounced or positive) and positive scores (vs. less pronounced or negative). Analyses were performed on data weighted for sex and educational level. *P*-values below <0.05 were considered significant (bold and underlined in table).
<sup>2</sup>In the 2022 survey, the questions regarding intention and vaccine uptake both had an extra answer option (‘not applicable’ and ‘don’t know’ respectively), which were excluded from analyses in order to ensure comparability of the surveys.
Proportions for intention are based on *n* = 965 for parents with a young child in 2022 and *n* = 912 for parents with an older child in 2022 and proportions for vaccine uptake are based on *n* = 993 for parents with a young child in 2022 and *n* = 995 for parents with an older child in 2022 (*n* obtained after weighting for sex and educational level).
<sup>3</sup>Items 1 and 4-7 concern beliefs about the effectiveness of vaccination, item 2 concerns beliefs about the safety of vaccination and item 3 concerns beliefs about the side effects of vaccination.

The proportion of parents who indicated that their child did not participate in the NIP was slightly but significantly larger in 2022 (Table 2). For all psychosocial factors of vaccine uptake, proportions of parents with negative scores were (slightly) greater in 2022 compared to 2013 (Table 2). In nearly all cases, these differences were significant: in 2022, significantly more parents had negative attitudes towards the NIP, negative beliefs about the effectiveness and side effects of vaccination, preferred natural infection over vaccination and reported lower trust towards the NIP (see Table 2 for the corresponding items). Likewise, for most psychosocial factors of vaccine uptake, proportions of parents with a positive score were smaller in 2022 compared to 2013 (Table 2). Differences in positive scores were less pronounced than differences in negative scores and significant in only two cases: in 2022, significantly less parents preferred natural infection over vaccination and felt that vaccinating their child is self-evident (Table 2).

For some of the psychosocial factors of vaccine uptake, both the proportion of parents with negative scores and the proportion of parents with positive scores were larger compared to 2013 (Table 2). In 2022, a significantly greater proportion of parents agreed that vaccinations could lead to severe side effects but at the same time a significantly greater proportion disagreed with that statement compared to parents in 2013. Other factors – attitudes, beliefs about the safety of vaccination and some of the beliefs about the effectiveness of vaccination (i.e., that vaccination is a good way to protect the health of adults and a good way to protect against severe complications of infectious diseases) – displayed similar trends but these results were not significant (Table 2).

All of the findings reported in the univariate analysis for differences in negative scores of psychosocial factors between parents in 2022 compared to 2013 held in the multivariate analysis (Table 3). That is, after controlling for demographic characteristics, it was shown that parents in 2022 had a significantly higher odds of negative attitudes towards the NIP, negative beliefs about the effectiveness and side effects of vaccination, a preference of natural infection over vaccination, and lower trust towards the NIP in 2022 compared to parents in 2013 (Table 3).

**Table 3.**
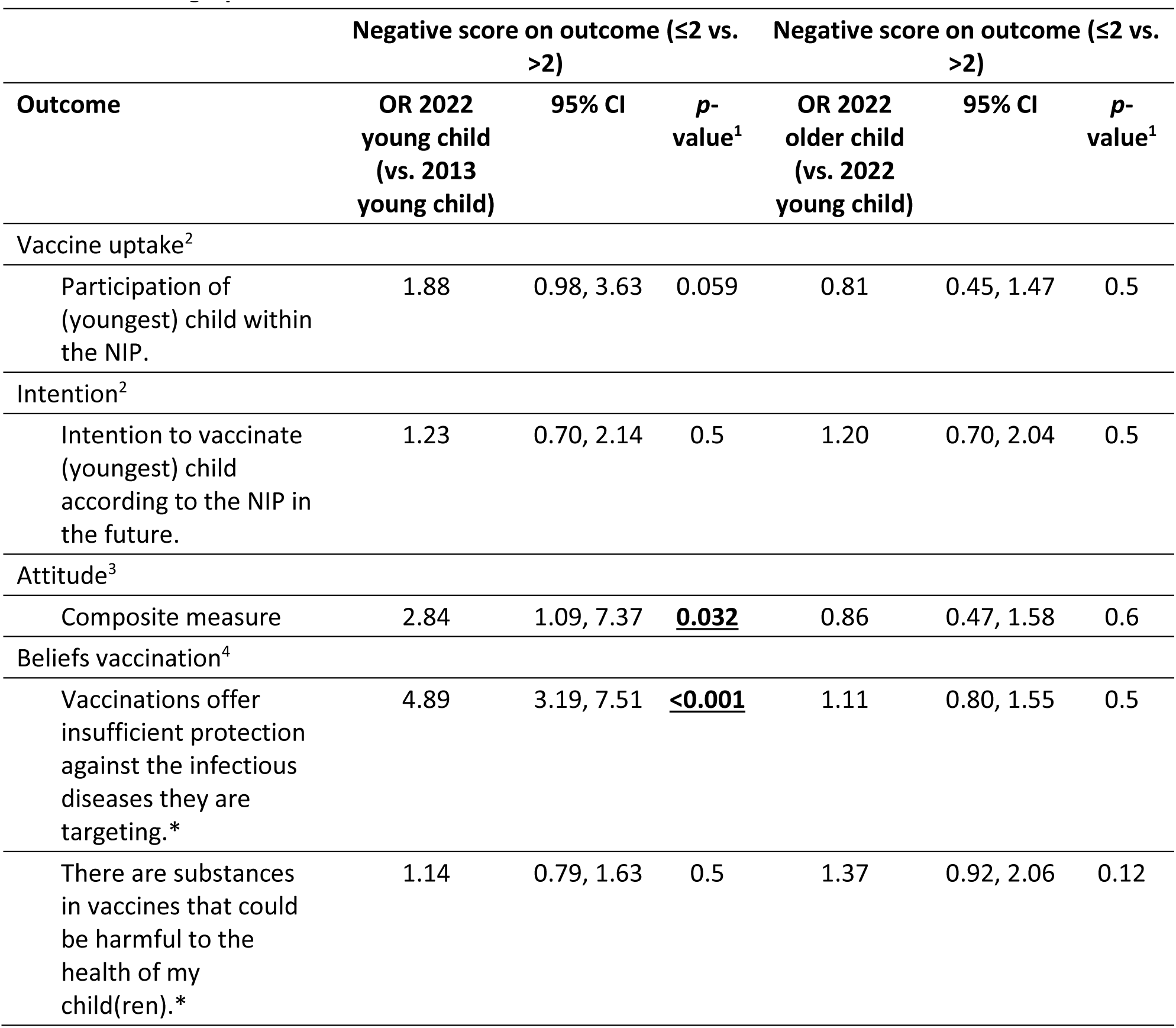
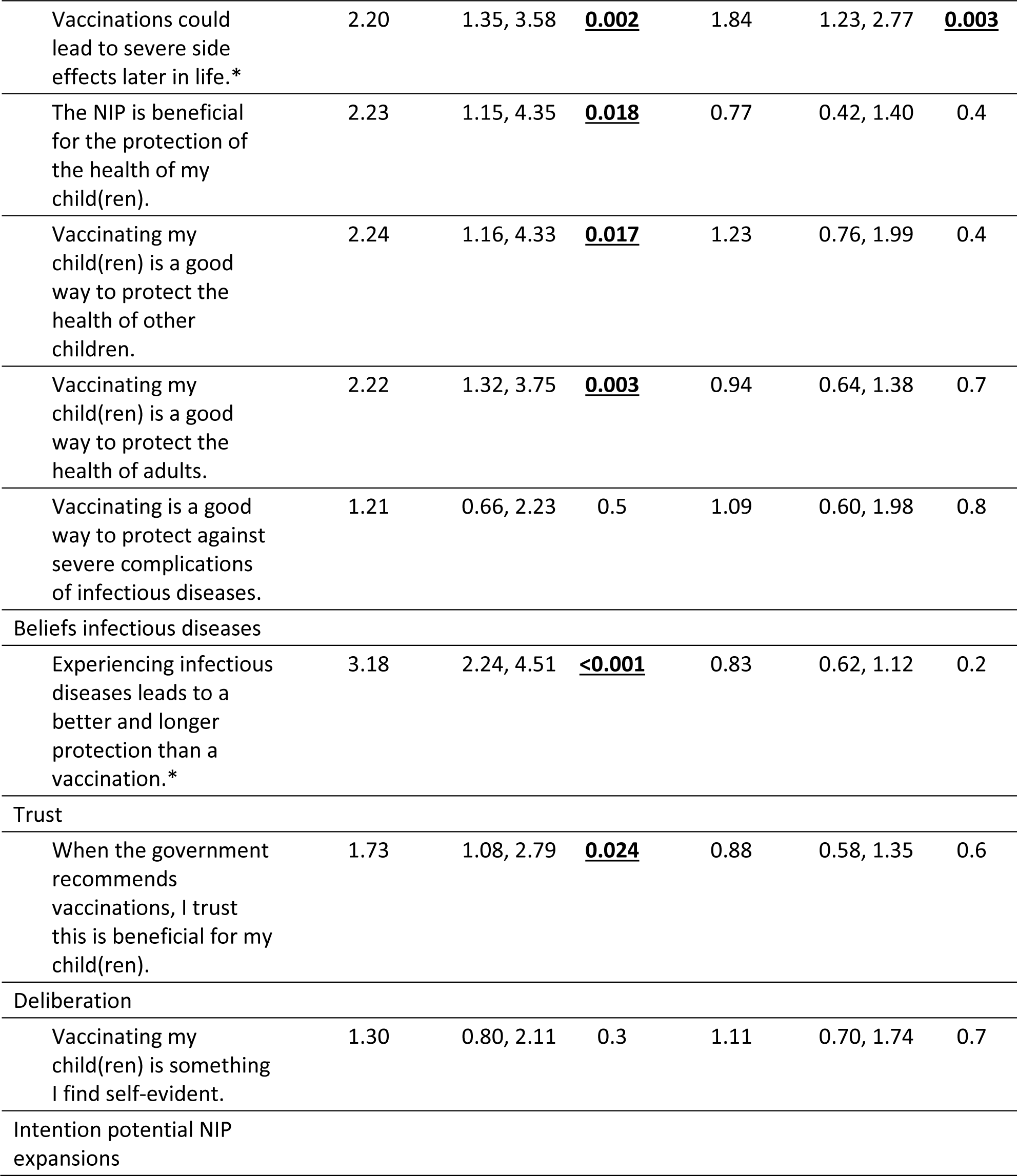
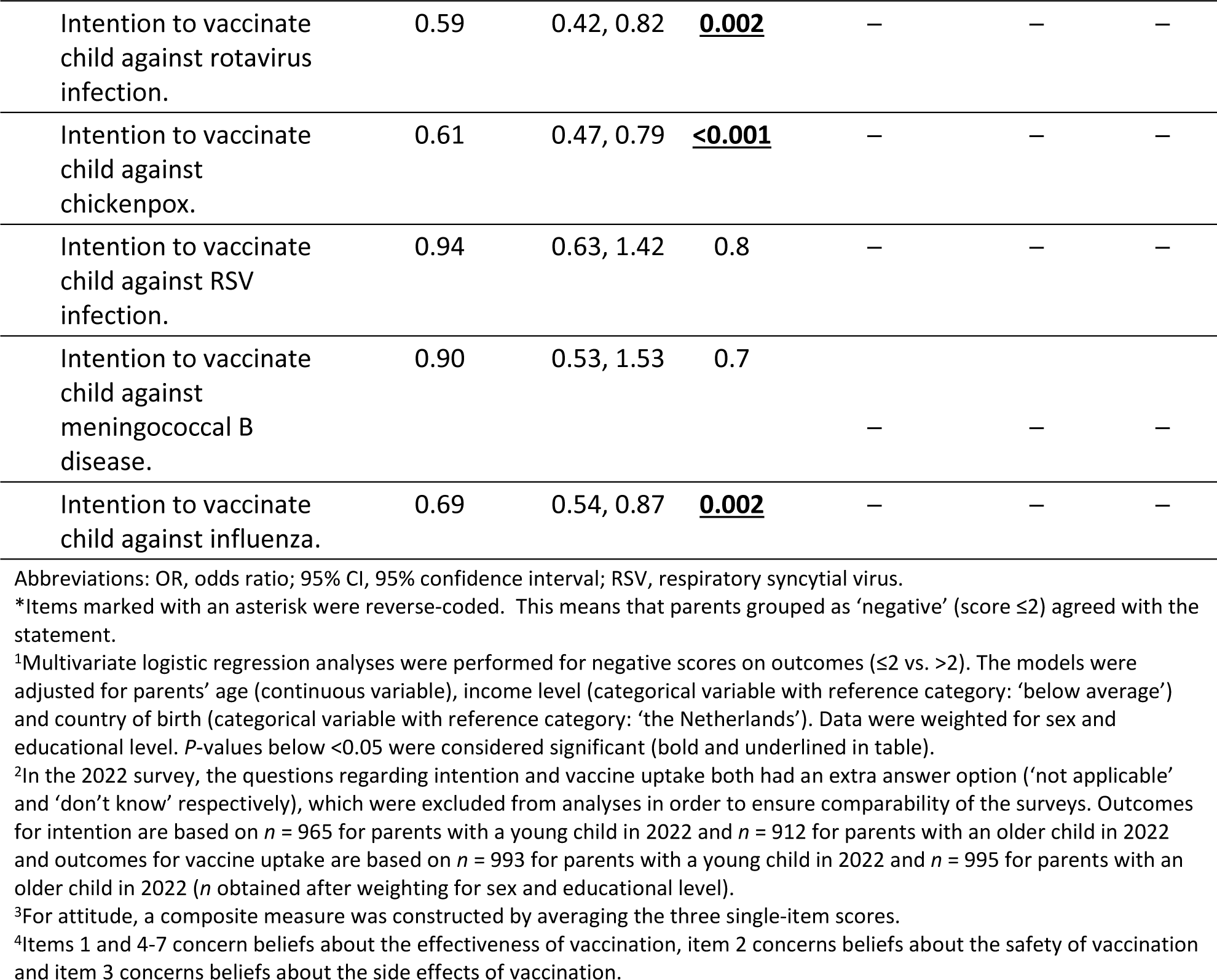
Multivariate comparison of negative vaccine uptake, intention, attitudes, beliefs, trust and deliberation towards the current NIP as well as negative intention towards potential expansions of the NIP between parents in 2013 and 2022. Data were weighted for sex and educational level and adjusted for other demographic characteristics.

#### Parents with an older child

Parents with an older child were largely similar to parents with a young child in 2022 in terms of their vaccine uptake, intention, attitudes, beliefs, trust and deliberation towards the NIP (Table 2, Table 3). Univariate analysis showed they only differed significantly from parents with a young child in the sense that significantly less parents with an older child preferred infection over vaccination and that significantly more of them did not believe that vaccinations offer insufficient protection or that vaccines contain harmful substances (Table 2). In the multivariate analysis they only differed significantly from parents with a young child in the sense that they had a higher odds of negative beliefs about the side effects of vaccination (Table 3).

### 3.3 Intention towards potential future expansions of the NIP

Generally, in 2022 parents with a young child were shown to be more positive in their intention towards potential future expansions of the NIP than parents in 2013 (Table 2, Table 3). Univariate analysis indicated that proportions of parents with a positive intention towards rotavirus infection, chickenpox, RSV infection, meningococcal B disease and influenza were significantly larger in 2022 (Table 2). Similarly, multivariate analysis showed that parents with a young child from 2022 had a significantly lower odds of a negative intention to vaccinate their child against rotavirus infection, chickenpox and influenza (Table 3).

### 3.4 Sensitivity analysis

The sensitivity analysis showed that aforementioned results largely held when different cut-off values were applied to the psychosocial factors of vaccine uptake (Supplemental Table 2, Supplemental Table 3).

## 4 Discussion

Different reports have shown that vaccine uptake within the Dutch NIP has been falling behind [6, 10]. This study suggests that most parents had positive intention, attitudes and trust towards the current NIP and perceived vaccinating their child to be self-evident in both 2022 and 2013. Scores for psychosocial factors among parents with an older child and parents with a young child in 2022 were similar. However, proportions of parents with negative scores were slightly larger in 2022 compared to 2013. In 2022, more parents tended to have a negative intention to vaccinate their child, negative attitudes about the value (good vs. bad), importance and necessity of vaccination, negative beliefs about the effectiveness, safety and side effects of vaccination, a preference of natural infection over vaccination and lower trust in the NIP. Parents in 2022 did not perceive vaccinating their child to be as self-evident as parents in 2013. Parents’ intention towards potential future expansions of the NIP, however, was more positive in 2022 compared to 2013.

Our results largely align with similar, recent studies from western countries. In Europe, there has been growing public concern towards the importance, effectiveness and safety of vaccinations in general [13, 14] and childhood vaccinations [15] since the COVID-19 pandemic. Furthermore, several studies from the USA [16–18] have suggested that, although parents’ attitudes towards the importance of vaccination and intention to vaccinate generally remained high during the COVID-19 pandemic, overall parental hesitancy towards childhood vaccination and concerns about the safety and side effects of vaccination increased. Insights from literature regarding parents’ beliefs about the effectiveness of vaccination were mixed: one study found that parental beliefs about the usefulness and effectiveness of vaccines remained stable and high [16], while another observed increasing concerns about vaccine efficacy much like we did [17]. Two other studies, on the other hand, presented different results. A study from the USA, carried out in an earlier stage of the pandemic, reported that parents had significantly lower negative attitudes towards childhood vaccination immediately after (vs. before) the onset of the pandemic, but that this effect disappeared by December 2020 [28]. A study among Canadian parents suggested that their perceptions towards the importance, safety and effectiveness of routine vaccination became more positive during the pandemic [29]. Finally, a study carried out in New-Zealand suggested that attitudes towards the safety of childhood vaccination were becoming increasingly polarised before the outbreak of the pandemic already [30]. For some of the psychosocial factors of vaccine uptake, our data also seemed to exhibit tendencies of polarisation. Our univariate analysis suggested that in 2022 significantly more parents had positive beliefs about the side effects of vaccination but also that significantly more parents had negative beliefs about side effects compared to 2013 (i.e., polarisation). Other factors – attitudes, beliefs about the safety of vaccination and some of the beliefs about the effectiveness of vaccination – also exhibited tendencies of polarisation but these results were not significant.

Our data show indications of more negative parental intention, attitudes, beliefs, trust and deliberation towards childhood vaccination in 2022. The substantial spread of (mis)information about vaccination during the pandemic may have played a role [31]. Previous studies have, for instance, noted the role of global scientific distrust [32], increased anti-vaccine searches on Google [33] and anti-vaccination sentiments on social media [34] in vaccine mistrust. Moreover, it has been suggested that parents’ beliefs about the effectiveness of routine childhood vaccines may have been affected by their perceptions of COVID-19 vaccines: while COVID-19 vaccines have been shown to be effective in protecting against hospitalisation and severe disease, they are less effective at preventing infection [35]. Furthermore, de Vries et al. [36] showed that many Dutch people held the belief that COVID-19 vaccination was the only solution to end the COVID-19 crisis at the beginning of 2021. However, during the winter of 2021 lockdown measures were implemented again, despite relatively high vaccination uptake rates [37]. De Vries et al. [36] have suggested that beliefs about the effectiveness of vaccination and vaccination acceptability may be affected by this initially low effectiveness of COVID-19 vaccination in preventing lockdown measures. We speculate that this may perhaps have influenced perceptions and trust towards childhood vaccination as well. Finally, the pandemic has exerted pressure on individuals to get vaccinated, partly driven by the [implementation of COVID-19 access passes](https://zoek.officielebekendmakingen.nl/stcrt-2021-33032.html). Our speculation is that this pressure could have shaped attitudes towards vaccination and impacted trust in the authorities involved.

Perceived barriers may have influenced parents’ vaccination decisions as well. Barriers are often linked to the intention-behaviour gap, but in the study conducted by Harmsen [21], it was suggested that high perceived barriers could also have a negative effect on parents’ intention to vaccinate their child. In 2022, child health clinic professionals indicated that parents seemed to suffer from “vaccine fatigue” [19] and felt too many vaccinations were being offered in the same period of time. The COVID-19 vaccinations scheduled during the second half of 2021 for teenagers aged 12-17 years [38] and during the spring of 2022 for children aged 5-11 years [39] were primarily scheduled alongside NIP group vaccinations offered to teenagers and children in those same age groups. This may have affected parents’ willingness to vaccinate.

Another possibility is that parents’ intention, attitudes, beliefs, trust and deliberation towards vaccination had already declined prior to the COVID-19 pandemic. The aforementioned study from New-Zealand already suggested a growing polarisation in attitudes regarding the safety of childhood vaccination before the pandemic’s onset [30]. In the Netherlands, there has been a decrease in vaccine uptake within the NIP since reporting year 2015 already (albeit with a short revival in the 2 years before the COVID-19 pandemic and another significant decline during the COVID-19 pandemic) and in 2019 the World Health Organisation (WHO) declared vaccine hesitancy to be one of the top ten major threats to global health [7]. Previous studies have, for instance, noted a trend of declining trust in public institutions within western populations long before the pandemic [40] and have linked this to increased vaccine hesitancy [41].

In contrast to our findings concerning the current NIP, parental intention towards potential NIP expansions was shown to be more positive in 2022 compared to 2013. For some potential NIP candidates, such as vaccination against rotavirus infection, meningococcal B disease, chickenpox and RSV infection, this could be the result of increased media attention due to recent advisory reports of the Health Council of the Netherlands for vaccination against meningococcal disease in 2018, rotavirus in 2017 and 2021 and chickenpox in 2020, RSV re-emergence in the summer of 2021 due to low RSV exposure during the COVID-19 pandemic [42, 43] and the outbreak of meningococcal W disease in 2015-2018 [44, 45]. For others, reasons are not entirely clear but perhaps the COVID-19 pandemic has led to increased awareness about the existence of these candidate vaccines.

Several limitations need to be considered. First, the surveys’ response rates were low, especially the one for parents with a young child in 2022 (estimated response: 12.2%). This could indicate possible selection response. Although weighted data were representative of the Dutch population in terms of sex and educational level, they were generally not representative in terms of parents’ country of birth, household income level and region of residence. In 2022 and 2013, weighted data seemed to be underrepresented by parents with a low household income level (approximately 29.5% and 19.1% in the Dutch population in 2021^3^ and 2013 [46], respectively) and high household income level (approximately 63.3% and 65.7% in the Dutch population in 2021^3^ and 2013 [46], respectively). In 2022, weighted data were underrepresented by parents not born in the Netherlands (19.7% in the Dutch population aged 20-60 years in 2022 [47]). In 2013, weighted data were overrepresented by parents from the Southern part of the Netherlands (21.9% in the Dutch population aged 25-40 years in 2013 [48]) and underrepresented by parents from the western part of the Netherlands (48.4% in the Dutch population aged 25-40 years in 2013 [48]). We should also acknowledge that weighting does not necessarily ensure generalisability of the findings [49]. Second, in the comparison between surveys from 2013 and 2022 we corrected for certain important demographic differences between samples. However, residual confounding remains a possibility. Third, our study focused on intention, attitudes, beliefs, trust and deliberation but did not investigate other potential determinants of vaccine uptake, such as perceived barriers. Fourth, we may have been rather restrictive in determining the cut-off values in our analyses (negative: score ≤2 and positive: score ≥6). Our sensitivity analysis, nevertheless, showed that results largely held when less restrictive cut-off values (negative: score ≤3 and positive: score ≥5) were applied to the psychosocial factors of vaccine uptake. Finally, due to the relatively long time period between the surveys, it is difficult to establish a causal relationship between the COVID-19 pandemic and the observed changes in parental perspectives towards childhood vaccination.

## 5 Conclusion

This study showed that most parents had positive intention, attitudes and trust towards childhood vaccination and perceived vaccinating their child to be self-evident in 2022. Simultaneously, proportions of parents with negative scores were slightly larger in 2022 compared to 2013. In 2022, more parents tended to have a negative intention to vaccinate their child, negative attitudes towards vaccination, negative beliefs about the effectiveness, safety and side effects of vaccination, a preference of natural infection over vaccination and lower trust in the NIP. Parents in 2022 did not perceive vaccinating their child to be as self-evident as parents in 2013 did. Monitoring these determinants of vaccine uptake and developing appropriate interventions could contribute to sustaining high vaccine uptake.

## Data Availability

All data produced in the present study are available in aggregated and anonymized form upon reasonable request to the authors.

## Author contributions

AL, JMH, HM were involved in conception and design of the study; AL, HM coordinated the study; MK, AL, HM interpreted and analysed the data and drafted the manuscript; MB collaborated in the statistical analysis; MK, AL, HM, JMH, MB, MV contributed to the interpretation of the results, critically reviewed and approved the final manuscript. All authors attest they meet the ICMJE criteria for authorship.

## Acknowledgements

We would like to thank Flycatcher Internet Research for conducting the data collection and all survey respondents for their participation. Furthermore, we would like to thank Christiaan Oostdijk for his contribution to the conception and design of the study.

## Footnotes

1 Parents with a young child were approached by two of Flycatcher’s partner agencies. One of the partner agencies sent out 3,308 invitations via email. The other partner agency did not send out email invitations but rather invited parents with a young child to participate in the survey when they logged on to their website. On January 13^th^, 2023, there were 5,000 parents with a young child in their panel. In the end, Flycatcher excluded 119 parents who did not have a young child after all.

2 In total, 4,837 invitations were sent out, but 58 parents did not have an older child after all and were excluded from further analyses by Flycatcher.

3 Information regarding household income was not yet available for 2022 from Statistics Netherlands. Data for 2021 are provisional.

