## Supplementary material for "Parental intention, attitudes, beliefs, trust and deliberation towards childhood vaccination in the Netherlands in 2022: Indications of change compared to 2013"

*Supplemental Table 1.* **Overview of questions from the 2022 survey and corresponding questions from the 2013 survey.**

| **Concept** | **Question type** | **2013 survey** | **2022 survey** |
|---|---|---|---|
| Age[1] | Open-ended | What is your date of birth?<br>- Day-Month-Year | What is your age?<br>- … years |
| Sex[1] | Multiple-choice | What is your sex?<br>o male<br>o female | What is your sex?<br>o female<br>o male<br>o other |
| Education[1] | | What is your highest completed level of education? (apart from primary education this means you have received a degree for said level of education)<br>o low (none, primary, lower vocational or lower secondary education)<br>o medium (secondary vocational or higher secondary education)<br>o high (higher professional or university education) | What is the highest level of education you have completed? (apart from primary education this means you have received a degree for said level of education)<br>o low (none, primary, lower vocational or lower secondary education)<br>o medium (secondary vocational or higher secondary education)<br>o high (higher professional or university education) |
| Income[1] | Multiple-choice | What is the gross annual income of your household? (this is the gross annual salary of all members of the household, including holiday allowance and 13 month's salary)<br>o minimum (less than €11,000)<br>o below average (between €11,000 and €23,000)<br>o average (between €23,000 and €34,000)<br>o 1 to 2 times average (between €34,000 and €56,000)<br>o 2 times average or more (€56,000 or more)<br>o don't know/prefer not to say | What is the gross annual income of your household? (this is the gross annual salary of all members of the household, including holiday allowance and 13 month's salary)<br>o minimum (less than €14,100)<br>o below average (between €14,100 and €36,500)<br>o average (between €36,500 and €43,500)<br>o 1 to 2 times average (between €43,500 and €73,000)<br>o 2 times average or more (€73,000 or more)<br>o don't know/prefer not to say |

*Supplemental Table 1.* **Overview of questions from the 2022 survey and corresponding questions from the 2013 survey.**

| **Concept** | **Question type** | **2013 survey** | **2022 survey** |
|---|---|---|---|
| Country of birth[1] | Multiple-choice (with 1 open-ended option) | What is your country of birth?<br>o the Netherlands<br>o Suriname<br>o (former) Netherlands Antilles and Aruba<br>o Turkey<br>o Morocco<br>o other, namely: … | What is your country of birth?<br>o the Netherlands<br>o other, namely: … |
| Other parent's/guardian's country of birth | Multiple-choice (with 1 open-ended option) | What is the country of birth of the other parent of your (youngest) child?<br>o the Netherlands<br>o Suriname<br>o (former) Netherlands Antilles and Aruba<br>o Turkey<br>o Morocco<br>o other, namely: …<br>o don't know | What is the country of birth of the other parent/guardian of your child?<br>o the Netherlands<br>o other, namely: …<br>o don't know/not applicable |
| Residence[1] | 2013: open-ended;<br>2022: multiple-choice | What is your postal code?<br>… | In which province do you live?<br>o Drenthe<br>o Flevoland<br>o Friesland<br>o Gelderland<br>o Groningen<br>o Limburg<br>o Noord-Brabant<br>o Noord-Holland<br>o Overijssel<br>o Utrecht<br>o Zeeland<br>o Zuid-Holland |

***Supplemental Table 1.*** **Overview of questions from the 2022 survey and corresponding questions from the 2013 survey.**

| **Concept** | **Question type** | **2013 survey** | **2022 survey** |
|---|---|---|---|
| Child in target age group | | *(not available)* | Do you have a child younger than 3.5 years?[2]<br>o yes<br>o no *(in case they answered 'no' the survey ended and no further questions were shown* |
| Number of children[1] | Open-ended | *(see next question)* | How many children do you have <u>in total</u>?<br>- … |
| Age of children[1] | Open-ended | Could you indicate the date of birth of all your children living at home?<br>- child 1: Day-Month-Year<br>- child 2: Day-Month-Year<br>- child 3: Day-Month-Year<br>- child 4: Day-Month-Year<br>*(etc.)* | How old is/are your child(ren)?<br>- … years *(number of options was dependent on number of children)* |
| Vaccine uptake | Multiple-choice | Is your youngest child vaccinated according to the national immunisation programme (NIP)?<br>o yes, fully (my youngest child has received all vaccinations they should have given their age)<br>o yes, partially (my youngest child has received some vaccinations they should have given their age but not others)<br>o no, not at all (my youngest child has not received any of the vaccinations they should have given their age) | Does your (youngest) child participate within the National Immunisation Programme?<br>With 'youngest child' we refer to your youngest child within the age group up to 3.5 years.[3]<br>o yes, they have received all vaccinations they should have given their age<br>o yes, they have received some vaccinations but not others<br>o no<br>o don't know |
| | | What is the reason your child is not/partially vaccinated? (multiple answers possible)<br>o my child was ill and could therefore not get vaccinated | What is/are the reason(s) your (youngest) child is not vaccinated according to the National Immunisation Programme? (multiple answers |

***Supplemental Table 1.*** **Overview of questions from the 2022 survey and corresponding questions from the 2013 survey.**

| Concept | Question type | 2013 survey | 2022 survey |
|---|---|---|---|
| | | o did not manage to make an appointment in time<br>o because I had doubts<br>o other, namely: … | possible) *(only shown to respondents who answered 'no' to the previous question)*<br>o my (youngest) child was ill and could therefore not get vaccinated<br>o did not manage to make an appointment in time[4]<br>o advice from a child health clinic professional[5]<br>o because I had doubts about vaccinating my (youngest) child<br>o my (youngest) child is healthy/has a strong immune system<br>o religious beliefs<br>o anthroposophical beliefs<br>o homeopathic beliefs<br>o vaccines/vaccinations are not halal<br>o I am against vaccinations<br>o I am against certain vaccine(s)<br>o people around me do not vaccinate their child(ren)<br>o negative messages about vaccines/vaccination in the news or on social media<br>o no trust in the National Immunisation Programme<br>o no trust in the government<br>o afraid of side effects<br>o bad experience with vaccines/vaccination<br>o due to COVID-19 measures my appointment at the child health clinic was canceled[5] |

*Supplemental Table 1.* **Overview of questions from the 2022 survey and corresponding questions from the 2013 survey.**

| **Concept** | **Question type** | **2013 survey** | **2022 survey** |
|---|---|---|---|
| | | | o due to COVID-19 measures I had to go to a different location<br>o I was afraid I myself and/or my (youngest) child would get infected with the COVID-19 virus at the child health clinic[5]<br>o I was afraid my (youngest) child would receive COVID-19 vaccinations without my knowledge<br>o due to COVID-19 I do not want my (youngest) child to get vaccinated anymore<br>o I do not want my data to be known by the RIVM<br>o other, namely: … |
| | | *(see previous question)* | What is/are the reason(s) your (youngest) child did receive some of the vaccinations within the National Immunisation Programme but not others? (multiple answers possible) *(only shown to respondents who answered 'yes, they have received some vaccinations but not others' to the previous question)*<br>- *(answer options were identical to those of the previous question)* |
| Intention | 7-point Likert scale | Do you intend to vaccinate your child(ren) according to the NIP in the future?<br>- 1 = certainly not to 7 = certainly | Do you intend to vaccinate your (youngest) child according to the National Immunisation Programme (NIP) in the future?<br>- 1 = certainly not to 7 = certainly (also including option 'not applicable, has already received all NIP vaccinations') |

*Supplemental Table 1.* **Overview of questions from the 2022 survey and corresponding questions from the 2013 survey.**

| Concept | Question type | 2013 survey | 2022 survey |
|---|---|---|---|
| Attitude | 7-point Likert scale | I think vaccinating my child(ren) within the NIP is:<br>1) 1 = very bad to 7 = very good | I think vaccinating my child(ren) according to the National Immunisation Programme is:<br>1) 1 = very bad to 7 = very good |
| | | 2) 1 = very unimportant to 7 = very important | 2) 1 = very unimportant to 7 = very important |
| | | 3) 1 = very unnecessary to 7 = very necessary | 3) 1 = very unnecessary to 7 = very necessary |
| Beliefs vaccination | 7-point Likert scale | To what extent do you agree with the following statements?<br>1) Vaccinations within the NIP offer insufficient protection against the infectious diseases they are targeting.<br>- 1 = completely disagree to 7 = completely agree | To what extent do you agree or disagree with the following statements? With vaccinations we refer to vaccinations within the National Immunisation Programme intended for your child(ren). We do not refer to COVID-19 vaccinations.<br>1) Vaccinations offer insufficient protection against the infectious diseases they are targeting.<br>- 1 = completely disagree to 7 = completely agree |
| | | 2) There are substances in vaccinations within the NIP that could be harmful to the health of my child.<br>- 1 = completely disagree to 7 = completely agree | 2) There are substances in vaccines that could be harmful to the health of my child(ren).<br>- 1 = completely disagree to 7 = completely agree |
| | | 3) Vaccinations within the NIP could lead to severe side effects later in life.<br>- 1 = completely disagree to 7 = completely agree | 3) Vaccinations could lead to severe side effects later in life.<br>- 1 = completely disagree to 7 = completely agree |
| | | 4) The NIP is beneficial for the protection of the health of my child. | 4) The National Immunisation Programme is beneficial for the protection of the health of my child(ren). |

*Supplemental Table 1.* **Overview of questions from the 2022 survey and corresponding questions from the 2013 survey.**

| Concept | Question type | 2013 survey | 2022 survey |
|---|---|---|---|
| | | - 1 = completely disagree to 7 = completely agree | - 1 = completely disagree to 7 = completely agree |
| | | 5) Vaccinating my child(ren) within the NIP is a good way to protect the health of children in the surroundings of my child(ren).<br>- 1 = completely disagree to 7 = completely agree | 5) Vaccinating my child(ren) is a good way to protect the health of other children.<br>- 1 = completely disagree to 7 = completely agree |
| | | 6) Vaccinating my child(ren) within the NIP is a good way to protect the health of adults in the surroundings of my child(ren).<br>- 1 = completely disagree to 7 = completely agree | 6) Vaccinating my child(ren) is a good way to protect the health of adults.<br>- 1 = completely disagree to 7 = completely agree |
| | | 7) By getting an infectious disease against which vaccinations are offered, complications may arise. Vaccinations within the NIP are a good way to protect against complications of infectious diseases.<br>- 1 = completely disagree to 7 = completely agree | 7) Vaccinating is a good way to protect against severe complications of infectious diseases.<br>- 1 = completely disagree to 7 = completely agree |
| Beliefs disease | 7-point Likert scale | To what extent do you agree with the following statements?<br>1) Experiencing infectious diseases leads to a better and lifelong protection than a vaccination.<br>- 1 = completely disagree to 7 = completely agree | To what extent do you agree or disagree with the following statements? With vaccinations we refer to vaccinations within the National Immunisation Programme intended for your child(ren). We do not refer to COVID-19 vaccinations.<br>1) Experiencing infectious diseases leads to a better and longer protection than a vaccination. |

***Supplemental Table 1.* Overview of questions from the 2022 survey and corresponding questions from the 2013 survey.**

| Concept | Question type | 2013 survey | 2022 survey |
|---|---|---|---|
| | | | - 1 = completely disagree to 7 = completely agree |
| Deliberation | 7-point Likert scale | To what extent do you agree with the following statements?<br>1) Vaccinating my child(ren) is something I find self-evident.<br>- 1 = completely disagree to 7 = completely agree | To what extent do you agree or disagree with the following statements? With vaccinations we refer to vaccinations within the National Immunisation Programme intended for your child(ren). We do not refer to COVID-19 vaccinations.<br>1) Vaccinating my child(ren) is something I find self-evident.<br>- 1 = completely disagree to 7 = completely agree |
| Trust | 7-point Likert scale | To what extent do you agree with the following statements?<br>1) When the government recommends vaccinations I trust this is beneficial for my child.<br>- 1 = completely disagree to 7 = completely agree | To what extent do you agree or disagree with the following statements? With vaccinations we refer to vaccinations within the National Immunisation Programme intended for your child(ren). We do not refer to COVID-19 vaccinations.<br>1) When the government recommends vaccinations, I trust this is beneficial for my child(ren).<br>- 1 = completely disagree to 7 = completely agree |
| | | *(not available)* | 2) I trust the vaccinations that are used within the National Immunisation Programme.<br>- 1 = completely disagree to 7 = completely agree |
| Intention potential NIP expansions | 7-point Likert scale | Hereunder a few diseases or pathogens are mentioned against which vaccinations might be offered within the NIP in the future. If you were | Would you vaccinate your child against the following infectious diseases if these vaccinations are incorporated within the National |

***Supplemental Table 1.*** **Overview of questions from the 2022 survey and corresponding questions from the 2013 survey.**

| **Concept** | **Question type** | **2013 survey** | **2022 survey** |
|---|---|---|---|
| | | provided with the choice of vaccinating your child, would you vaccinate your child against …<br>1) Rotavirus (severe diarrhoea in children)<br>- 1 = certainly not to 7 = certainly | Immunisation Programme? (all vaccinations within the National Immunisation Programme are free of charge) *(this question was only shown to parents with at least one child aged <3.5 years)*<br>1) Rotavirus infection (severe diarrhoea in children); this concerns an oral vaccine for which an injection is not necessary<br>- 1 = certainly not to 7 = certainly |
| | | 2) Chickenpox<br>- 1 = certainly not to 7 = certainly | 2) Chickenpox<br>- 1 = certainly not to 7 = certainly |
| | | 3) RSV (respiratory syncytial virus, severe form of breathlessness in babies)<br>- 1 = certainly not to 7 = certainly | 3) RSV infection (respiratory syncytial virus, severe form of breathlessness in babies)<br>- 1 = certainly not to 7 = certainly |
| | | 4) Meningococcal B<br>- 1 = certainly not to 7 = certainly | 4) Meningococcal B disease (sepsis or meningitis)<br>- 1 = certainly not to 7 = certainly |
| | | 5) Influenza<br>- 1 = certainly not to 7 = certainly | 5) Influenza<br>- 1 = certainly not to 7 = certainly |
| | | | 6) Hepatitis A (liver infection: jaundice, fatigue, slight fever, sometimes upper abdominal pain and nausea)<br>- 1 = certainly not to 7 = certainly |
| Opinion NIP invitations | 5-point Likert scale | *(not available)* | What is your opinion on the invitations for the vaccinations within the National Immunisation Programme?<br>o very clear |

*Supplemental Table 1.* **Overview of questions from the 2022 survey and corresponding questions from the 2013 survey.**

| **Concept** | **Question type** | **2013 survey** | **2022 survey** |
|---|---|---|---|
| | | | o a little clear<br>o not clear/not unclear<br>o not very clear<br>o not clear at all<br>o don't know, as I did not receive the invitation(s)<br>o don't know, I did not open the invitation(s)<br>o no opinion |
| | Open-ended | *(not available)* | Could you indicate what you do not find clear about the invitation(s)? *(only shown to respondents who answered 'not clear at all' or 'not very clear' to the previous question)*<br>- … |
| Informed consent | Multiple-choice | *(not available)* | Please first read the provided explanation. If your child receives a vaccination, would you give permission to pass on data to the RIVM? This concerns your child's personal data along with their vaccination data.<br>o yes<br>o no<br>o don't know/no opinion |
| | Open-ended | *(not available)* | Why would you not give permission to pass on data to the RIVM? *(only shown to respondents who answered 'no' to the previous question)*<br>- … |
| Opinion NIP | 5-point Likert scale | *(not available)* | Has your opinion about the National Immunisation Programme changed because |

*Supplemental Table 1.* **Overview of questions from the 2022 survey and corresponding questions from the 2013 survey.**

| **Concept** | **Question type** | **2013 survey** | **2022 survey** |
|---|---|---|---|
| | | | permission is being asked to pass on data to the RIVM? My opinion has become:<br>- 1 = much more positive to 5 = much more negative |
| | | Has your opinion about vaccinations changed since you have made the decision to vaccinate or not vaccinate your oldest child?<br>- 1 = much more positive to 5 = much more negative | Has your opinion about vaccinations within the National Immunisation Programme changed since you have or have not vaccinated your (oldest) child? My opinion has become<br>- 1 = much more positive to 5 = much more negative: |
| | | *(not available)* | Has your opinion about vaccinations within the National Immunisation Programme changed because of the COVID-19 pandemic? My opinion has become:<br>- 1 = much more positive to 5 = much more negative |
| Other | Open-ended | In case you have any comments as a result of this questionnaire, feel free to use the space below.<br>- … | In case you have any comments about the subject of this questionnaire, feel free to use the space below.<br>- … |

[1]In 2013, respondents from another agency's panel were invited in addition to the Flycatcher panel in order to obtain sufficient responses from immigrant parents. Questions concerning sex, age, country of birth, residence, income, education and their children's date of birth were only shown to the parents from the additional panel and not to the Flycatcher panel as this information was already available for the latter. The question concerning the other parent's/guardian's country of birth was shown to both the Flycatcher panel and the additional panel.

[2]To the target group of parents assumed to have at least one child aged 9-14 years, the question "Do you have a child between 9 and 14 years?" was shown rather than "Do you have a child younger than 3.5 years?".

[3]To the target group of parents assumed to have at least one child aged 9-14 years, the statement "With 'youngest child' we refer to your youngest child within the age group between 9 and 14 years." was shown rather than "With 'youngest child' we refer to your youngest child within the age group up to 3.5 years.".

[4]To the target group of parents with at least one child aged 9-14 years, the answer option that was provided was "the date/time at which the vaccination was offered did not suit me" rather than "did not manage to make an appointment in time".

***Supplemental Table 1.*** **Overview of questions from the 2022 survey and corresponding questions from the 2013 survey.**

| Concept | Question type | 2013 survey | 2022 survey |
|---|---|---|---|

[5]The survey for parents with at least one child aged 9-14 years referred to “vaccination location” rather than “child health clinic” and “youth health care professional” rather than “child health clinic professional”.

*Supplemental Table 2.* **Sensitivity analysis for the overview and univariate comparison of intention, attitudes, beliefs, trust and deliberation towards the current NIP and intention towards potential expansions of the NIP among parents in 2013 and 2022. Data were weighted for sex and educational level and different cut-off values were applied to the psychosocial factors of vaccine uptake.**

| | **2013** | **2022** | | ***p*-values[1]** | |
|---|---|---|---|---|---|
| | **Young child, *n* = 797** | **Young child, *n* = 998** | **Older child, *n* = 1002** | **2013 vs. 2022 young child** | **2022 young child vs. older child** |
| *Intention[2]* | | | | | |
| Intention to vaccinate (youngest) child according to the NIP in the future. | | | | | |
| Negative (score 1-3) | 6.4% | 7.1% | 8.7% | 0.7 | 0.2 |
| Less pronounced (score 4) | 3.6% | 2.3% | 3.8% | | |
| Positive (score 5-7) | 90.0% | 90.6% | 87.5% | 0.7 | **<u>0.037</u>** |
| *Attitude* | | | | | |
| I think vaccinating my child(ren) according to the NIP is: very bad – very good | | | | | |
| Negative (score 1-3) | 3.3% | 6.5% | 6.0% | **<u>0.007</u>** | 0.6 |
| Less pronounced (score 4) | 10.5% | 10.2% | 9.0% | | |
| Positive (score 5-7) | 86.2% | 83.2% | 85.0% | 0.2 | 0.3 |
| I think vaccinating my child(ren) according to the NIP is: very unimportant – very important | | | | | |
| Negative (score 1-3) | 3.9% | 6.7% | 6.3% | 0.063 | 0.7 |
| Less pronounced (score 4) | 10.2% | 8.8% | 9.0% | | |
| Positive (score 5-7) | 85.9% | 84.5% | 84.8% | 0.5 | 0.9 |
| I think vaccinating my child(ren) according to the NIP is: very unnecessary – very necessary | | | | | |
| Negative (score 1-3) | 5.9% | 9.6% | 8.4% | **<u>0.012</u>** | 0.3 |
| Less pronounced (score 4) | 12.9% | 10.1% | 10.3% | | |
| Positive (score 5-7) | 81.2% | 80.3% | 81.4% | 0.7 | 0.6 |
| *Beliefs vaccination[3]* | | | | | |
| Vaccinations offer insufficient protection against the infectious diseases they are targeting.* | | | | | |
| Negative (score 1-3) | 10.4% | 33.8% | 27.1% | **<u><0.001</u>** | **<u>0.002</u>** |
| Less pronounced (score 4) | 30.2% | 13.8% | 12.9% | | |
| Positive (score 5-7) | 59.4% | 52.5% | 60.0% | **<u>0.018</u>** | **<u>0.001</u>** |

*Supplemental Table 2.* **Sensitivity analysis for the overview and univariate comparison of intention, attitudes, beliefs, trust and deliberation towards the current NIP and intention towards potential expansions of the NIP among parents in 2013 and 2022. Data were weighted for sex and educational level and different cut-off values were applied to the psychosocial factors of vaccine uptake.**

| | **2013** | **2022** | | ***p*-values[1]** | |
|---|---|---|---|---|---|
| | **Young child, *n* = 797** | **Young child, *n* = 998** | **Older child, *n* = 1002** | **2013 vs. 2022 young child** | **2022 young child vs. older child** |
| There are substances in vaccines that could be harmful to the health of my child(ren).* | | | | | |
| Negative (score 1-3) | 36.9% | 26.4% | 23.3% | **<u><0.001</u>** | 0.13 |
| Less pronounced (score 4) | 42.2% | 26.3% | 25.4% | | |
| Positive (score 5-7) | 20.9% | 47.3% | 51.2% | **<u><0.001</u>** | 0.092 |
| Vaccinations could lead to severe side effects later in life.* | | | | | |
| Negative (score 1-3) | 16.3% | 21.8% | 20.6% | **<u>0.02</u>** | 0.5 |
| Less pronounced (score 4) | 47.2% | 21.2% | 23.7% | | |
| Positive (score 5-7) | 36.5% | 56.9% | 55.7% | **<u><0.001</u>** | 0.6 |
| The NIP is beneficial for the protection of the health of my child(ren). | | | | | |
| Negative (score 1-3) | 3.6% | 8.9% | 7.6% | **<u><0.001</u>** | 0.3 |
| Less pronounced (score 4) | 14.5% | 12.8% | 12.0% | | |
| Positive (score 5-7) | 81.8% | 78.3% | 80.5% | 0.14 | 0.3 |
| Vaccinating my child(ren) is a good way to protect the health of other children. | | | | | |
| Negative (score 1-3) | 4.5% | 10.3% | 10.3% | **<u><0.001</u>** | >0.9 |
| Less pronounced (score 4) | 17.3% | 11.6% | 14.0% | | |
| Positive (score 5-7) | 78.2% | 78.0% | 75.7% | >0.9 | 0.2 |
| Vaccinating my child(ren) is a good way to protect the health of adults. | | | | | |
| Negative (score 1-3) | 10.0% | 13.6% | 12.6% | 0.067 | 0.5 |
| Less pronounced (score 4) | 23.8% | 18.1% | 18.8% | | |
| Positive (score 5-7) | 66.2% | 68.3% | 68.6% | 0.4 | 0.9 |
| Vaccinating is a good way to protect against severe complications of infectious diseases. | | | | | |
| Negative (score 1-3) | 5.7% | 8.5% | 6.0% | 0.08 | **<u>0.034</u>** |
| Less pronounced (score 4) | 23.4% | 12.3% | 12.3% | | |

*Supplemental Table 2.* **Sensitivity analysis for the overview and univariate comparison of intention, attitudes, beliefs, trust and deliberation towards the current NIP and intention towards potential expansions of the NIP among parents in 2013 and 2022. Data were weighted for sex and educational level and different cut-off values were applied to the psychosocial factors of vaccine uptake.**

| | **2013** | **2022** | | ***p*-values[1]** | |
|---|---|---|---|---|---|
| | **Young child, *n* = 797** | **Young child, *n* = 998** | **Older child, *n* = 1002** | **2013 vs. 2022 young child** | **2022 young child vs. older child** |
| Positive (score 5-7) | 70.8% | 79.1% | 81.7% | **<u>0.001</u>** | 0.2 |
| *Beliefs disease* | | | | | |
| Experiencing infectious diseases leads to a better and longer protection than a vaccination.* | | | | | |
| Negative (score 1-3) | 28.5% | 41.5% | 37.1% | **<u><0.001</u>** | 0.056 |
| Less pronounced (score 4) | 31.4% | 29.9% | 35.3% | | |
| Positive (score 5-7) | 40.1% | 28.6% | 27.6% | **<u><0.001</u>** | 0.6 |
| *Trust* | | | | | |
| When the government recommends vaccinations, I trust this is beneficial for my child(ren). | | | | | |
| Negative (score 1-3) | 11.2% | 13.1% | 12.3% | 0.3 | 0.6 |
| Less pronounced (score 4) | 15.8% | 15.4% | 14.7% | | |
| Positive (score 5-7) | 73.0% | 71.5% | 72.9% | 0.6 | 0.5 |
| *Deliberation* | | | | | |
| Vaccinating my child(ren) is something I find self-evident. | | | | | |
| Negative (score 1-3) | 9.1% | 11.2% | 11.8% | 0.2 | 0.7 |
| Less pronounced (score 4) | 9.9% | 13.2% | 11.8% | | |
| Positive (score 5-7) | 81.0% | 75.6% | 76.4% | **<u>0.032</u>** | 0.7 |
| *Intention potential NIP expansions* | | | | | |
| Intention to vaccinate child against rotavirus infection. | | | | | |
| Negative (score 1-3) | 25.6% | 16.8% | – | **<u><0.001</u>** | – |
| Less pronounced (score 4) | 23.8% | 20.6% | – | | |
| Positive (score 5-7) | 50.6% | 62.6% | – | **<u><0.001</u>** | – |
| Intention to vaccinate child against chickenpox. | | | | | |
| Negative (score 1-3) | 34.4% | 26.3% | – | **<u>0.001</u>** | – |
| Less pronounced (score 4) | 18.4% | 16.9% | – | | |
| Positive (score 5-7) | 47.1% | 56.8% | – | **<u><0.001</u>** | – |

***Supplemental Table 2.*** **Sensitivity analysis for the overview and univariate comparison of intention, attitudes, beliefs, trust and deliberation towards the current NIP and intention towards potential expansions of the NIP among parents in 2013 and 2022. Data were weighted for sex and educational level and different cut-off values were applied to the psychosocial factors of vaccine uptake.**

| | **2013** | **2022** | | ***p*-values[1]** | |
|---|---|---|---|---|---|
| | **Young child, *n* = 797** | **Young child, *n* = 998** | **Older child, *n* = 1002** | **2013 vs. 2022 young child** | **2022 young child vs. older child** |
| Intention to vaccinate child against RSV infection. | | | | | |
| Negative (score 1-3) | 12.1% | 11.8% | – | 0.9 | – |
| Less pronounced (score 4) | 22.9% | 15.5% | – | | |
| Positive (score 5-7) | 65.1% | 72.7% | – | **<u>0.005</u>** | – |
| Intention to vaccinate child against meningococcal B disease. | | | | | |
| Negative (score 1-3) | 5.5% | 8.2% | – | 0.051 | – |
| Less pronounced (score 4) | 25.4% | 10.7% | – | | |
| Positive (score 5-7) | 69.1% | 81.1% | – | **<u><0.001</u>** | – |
| Intention to vaccinate child against influenza. | | | | | |
| Negative (score 1-3) | 54.7% | 49.2% | – | 0.059 | – |
| Less pronounced (score 4) | 19.7% | 19.6% | – | | |
| Positive (score 5-7) | 25.6% | 31.2% | – | **<u>0.039</u>** | – |
| Intention to vaccinate child against hepatitis A. | | | | | |
| Negative (score 1-3) | – | 12.0% | – | – | – |
| Less pronounced (score 4) | – | 15.1% | – | | |
| Positive (score 5-7) | – | 72.8% | – | – | – |

Abbreviations: IQR, interquartile range; SD, standard deviation; RSV, respiratory syncytial virus.

*Items marked with an asterisk were reverse-coded. This means that parents grouped as 'negative' (score 1-3) agreed with the statement and parents grouped as 'positive' (score 5-7) disagreed with the statement.

[1]Chi-square tests were used to compare negative (vs. less pronounced or positive) and positive scores (vs. less pronounced or negative). Analyses were performed on data weighted for sex and educational level. *P*-values below <0.05 were considered significant (bold and underlined in table).

[2]In the 2022 survey, the question regarding intention had an extra answer option ('not applicable'), which was excluded from analyses in order to ensure comparability of the surveys. Proportions for intention are based on *n* = 965 for parents with a young child in 2022 and *n* = 912 for parents with an older child in 2022 (*n* obtained after weighting for sex and educational level).

[3]Items 1 and 4-7 concern beliefs about the effectiveness of vaccination, item 2 concerns beliefs about the safety of vaccination and item 3 concerns beliefs about the side effects of vaccination.

***Supplemental Table 3.*** **Sensitivity analysis for the multivariate comparison of negative intention, attitudes, beliefs, trust and deliberation towards the current NIP as well as negative intention towards potential expansions of the NIP between parents in 2013 and 2022. Data were weighted for sex and educational level and adjusted for other demographic characteristics. Different cut-off values were applied to the psychosocial factors of vaccine uptake.**

| | **Negative score on outcome (≤3 vs. >3)** | | | **Negative score on outcome (≤3 vs. >3)** | | |
|---|---|---|---|---|---|---|
| **Outcome** | **OR 2022 young child (vs. 2013 young child)** | **95% CI** | ***p*-value[1]** | **OR 2022 older child (vs. 2022 young child)** | **95% CI** | ***p*-value[1]** |
| Intention[2] | | | | | | |
| Intention to vaccinate (youngest) child according to the NIP in the future. | 1.11 | 0.69, 1.79 | 0.7 | 1.29 | 0.81, 2.05 | 0.3 |
| Attitude[3] | | | | | | |
| Composite measure | 2.24 | 1.14, 4.42 | **0.020** | 0.98 | 0.60, 1.60 | >0.9 |
| Beliefs vaccination[4] | | | | | | |
| Vaccinations offer insufficient protection against the infectious diseases they are targeting.* | 4.68 | 3.41, 6.41 | **<0.001** | 0.93 | 0.71, 1.21 | 0.6 |
| There are substances in vaccines that could be harmful to the health of my child(ren).* | 0.60 | 0.47, 0.78 | **<0.001** | 1.31 | 0.97, 1.75 | 0.074 |
| Vaccinations could lead to severe side effects later in life.* | 1.64 | 1.20, 2.25 | **0.002** | 1.79 | 1.30, 2.47 | **<0.001** |
| The NIP is beneficial for the protection of the health of my child(ren). | 2.42 | 1.49, 3.93 | **<0.001** | 1.01 | 0.65, 1.56 | >0.9 |
| Vaccinating my child(ren) is a good way to protect the health of other children. | 2.66 | 1.59, 4.46 | **<0.001** | 1.13 | 0.76, 1.68 | 0.5 |
| Vaccinating my child(ren) is a good way to protect the health of adults. | 1.46 | 0.99, 2.17 | 0.057 | 1.00 | 0.72, 1.40 | >0.9 |
| Vaccinating is a good way to protect | 1.65 | 0.98, 2.80 | 0.060 | 0.82 | 0.52, 1.29 | 0.4 |

***Supplemental Table 3.*** **Sensitivity analysis for the multivariate comparison of negative intention, attitudes, beliefs, trust and deliberation towards the current NIP as well as negative intention towards potential expansions of the NIP between parents in 2013 and 2022. Data were weighted for sex and educational level and adjusted for other demographic characteristics. Different cut-off values were applied to the psychosocial factors of vaccine uptake.**

| | **Negative score on outcome (≤3 vs. >3)** | | | **Negative score on outcome (≤3 vs. >3)** | | |
|---|---|---|---|---|---|---|
| **Outcome** | **OR 2022 young child (vs. 2013 young child)** | **95% CI** | ***p*-value[1]** | **OR 2022 older child (vs. 2022 young child)** | **95% CI** | ***p*-value[1]** |
| against severe complications of infectious diseases. | | | | | | |
| Beliefs infectious diseases | | | | | | |
| Experiencing infectious diseases leads to a better and longer protection than a vaccination.* | 1.65 | 1.29, 2.11 | **<u><0.001</u>** | 0.87 | 0.68, 1.11 | 0.3 |
| Trust | | | | | | |
| When the government recommends vaccinations, I trust this is beneficial for my child(ren). | 1.22 | 0.83, 1.78 | 0.3 | 1.07 | 0.76, 1.52 | 0.7 |
| Deliberation | | | | | | |
| Vaccinating my child(ren) is something I find self-evident. | 1.31 | 0.88, 1.95 | 0.2 | 1.25 | 0.87, 1.79 | 0.2 |
| Intention potential NIP expansions | | | | | | |
| Intention to vaccinate child against rotavirus infection | 0.59 | 0.45, 0.79 | **<u><0.001</u>** | – | – | – |
| Intention to vaccinate child against chickenpox | 0.65 | 0.51, 0.83 | **<u><0.001</u>** | – | – | – |
| Intention to vaccinate child against RSV infection | 0.99 | 0.68, 1.44 | >0.9 | – | – | – |
| Intention to vaccinate child against | 1.36 | 0.85, 2.17 | 0.2 | – | – | – |

***Supplemental Table 3.*** **Sensitivity analysis for the multivariate comparison of negative intention, attitudes, beliefs, trust and deliberation towards the current NIP as well as negative intention towards potential expansions of the NIP between parents in 2013 and 2022. Data were weighted for sex and educational level and adjusted for other demographic characteristics. Different cut-off values were applied to the psychosocial factors of vaccine uptake.**

| | **Negative score on outcome (≤3 vs. >3)** | | | **Negative score on outcome (≤3 vs. >3)** | | |
|---|---|---|---|---|---|---|
| **Outcome** | **OR 2022 young child (vs. 2013 young child)** | **95% CI** | ***p*-value[1]** | **OR 2022 older child (vs. 2022 young child)** | **95% CI** | ***p*-value[1]** |
| meningococcal B disease | | | | | | |
| Intention to vaccinate child against influenza | 0.76 | 0.59, 0.96 | **0.022** | – | – | – |

Abbreviations: OR, odds ratio; 95% CI, 95% confidence interval; RSV, respiratory syncytial virus.
*Items marked with an asterisk were reverse-coded. This means that parents grouped as 'negative' (score ≤3) agreed with the statement.
[1]Multivariate logistic regression analyses were performed for negative scores on outcomes (≤3 vs. >3). The models were adjusted for parents' age (continuous variable), income level (categorical variable with reference category: 'below average') and country of birth (categorical variable with reference category: 'the Netherlands'). Data were weighted for sex and educational level. *P*-values below <0.05 were considered significant (bold and underlined in table).
[2]In the 2022 survey, the question regarding intention had an extra answer option ('not applicable'), which was excluded from analyses in order to ensure comparability of the surveys. Outcomes for intention are based on $n$ = 965 for parents with a young child in 2022 and $n$ = 912 for parents with an older child in 2022 ($n$ obtained after weighting for sex and educational level).
[3]For attitude, a composite measure was constructed by averaging the three single-item scores.
[4]Items 1 and 4-7 concern beliefs about the effectiveness of vaccination, item 2 concerns beliefs about the safety of vaccination and item 3 concerns beliefs about the side effects of vaccination.
